# Immunogenicity of an additional mRNA-1273 SARS-CoV-2 vaccination in people living with HIV with hyporesponse after primary vaccination

**DOI:** 10.1101/2022.08.10.22278577

**Authors:** Marlou J. Jongkees, Daryl Geers, Kathryn S. Hensley, Wesley Huisman, Corine H. GeurtsvanKessel, Susanne Bogers, Lennert Gommers, Grigorios Papageorgiou, Simon P. Jochems, Jan G. den Hollander, Emile F. Schippers, Heidi S.M. Ammerlaan, Wouter F.W. Bierman, Marc van der Valk, Marvin A.H. Berrevoets, Robert Soetekouw, Nienke Langebeek, Anke H.W. Bruns, Eliane M.S. Leyten, Kim C.E. Sigaloff, Marit G.A. van Vonderen, Corine E. Delsing, Judith Branger, Peter D. Katsikis, Yvonne M. Mueller, Rory D. de Vries, Bart J.A. Rijnders, Kees Brinkman, Casper Rokx, Anna H.E. Roukens

**Author notes:** **Corresponding author** Dr. Anna H.E. Roukens. Contributed equally to this manuscript (Daryl Geers, Kathryn S. Hensley, Casper Rokx, Anna H.E. Roukens).

## Abstract

**Background:** The COVIH study is a prospective SARS-CoV-2 vaccination study in people living with HIV (PLWH). Of the 1154 PLWH enrolled, 14% showed a reduced or absent antibody response after a primary vaccination regimen. As the response to an additional vaccination in PLWH with hyporesponse is unknown, we evaluated whether an additional vaccination boosts immune responses in these hyporesponders.

**Methods:** Consenting hyporesponders received an additional 100 µg of mRNA-1273. Hyporesponse was defined as ≤300 spike(S)-specific binding antibody units [BAU]/mL. The primary endpoint was the increase in antibodies 28 days after the additional vaccination. Secondary endpoints were the correlation between patient characteristics and antibody response, levels of neutralizing antibodies, S-specific T-cell and B-cell responses, and reactogenicity.

**Results:** Of the 75 PLWH enrolled, five were excluded as their antibody level had increased to >300 BAU/mL at baseline, two for a SARS-CoV-2 infection before the primary endpoint evaluation and two were lost to follow-up. Of the 66 remaining participants, 40 previously received ChAdOx1-S, 22 BNT162b2, and four Ad26.COV2.S. The median age was 63 [IQR:60-66], 86% were male, pre-vaccination and nadir CD4+ T-cell counts were 650/μL [IQR:423-941] and 230/μL [IQR:145-345] and 96% had HIV-RNA <50 copies/ml. The mean antibody level before the additional vaccination was 35 BAU/mL (SEM 5.4) and 45/66 (68%) were antibody negative. After the additional mRNA-1273 vaccination, antibodies were >300 BAU/mL in 64/66 (97%) with a mean increase of 4282 BAU/mL (95%CI:3241-5323). No patient characteristics correlated with the magnitude of the antibody response nor did the primary vaccination regimen. The additional vaccination significantly increased the proportion of participants with detectable ancestral S-specific B-cells (p=0.016) and CD4+ T-cells (p=0.037).

**Conclusion:** An additional mRNA-1273 vaccination induced a robust serological response in 97% of the PLWH with a hyporesponse after a primary vaccination regimen. This response was observed regardless of the primary vaccination regimen or patient characteristics.

## Introduction

People living with HIV (PLWH) show diminished responses to a wide variety of vaccines compared to HIV-negative controls, such as hepatitis B[1] and seasonal influenza vaccines[2]. As we hypothesized that this also holds true for SARS-CoV-2 vaccines, we previously investigated the immunogenicity of SARS-CoV-2 vaccinations in PLWH with the vaccines currently approved in the Netherlands. Whereas smaller studies showed variable antibody responses after SARS-CoV-2 vaccination in PLWH compared to HIV-negative controls[3–9], our study in 1154 PLWH clearly demonstrated a diminished antibody response[10]. In total, 165 (14.3%) of the participants were hyporesponders (≤300 spike (S)-specific binding antibody units [BAU]/mL by chemiluminescence immunoassay [DiaSorin Liaison]) and in 33 of them the antibody level remained below the cut-off level of test positivity (33.8 BAU/mL). In comparison with HIV-negative controls, hyporesponse rates were 6.7% (59/884) versus none (0/94) after vaccination with BNT162b2, 4.0% (4/100) versus none (0/247) after mRNA-1273, 54.7% (82/150) versus 38.5% (10/26) after ChAdOx1-S and all (20/20) versus 98.6% (72/73) after Ad26.COV2.S. CD4+ T-cell counts under 250 cells/µL, age above 65 years, and male sex were associated with lower antibody levels (all p≤0.001). The reduced response against SARS-CoV-2 in PLWH compared to HIV-negative controls is in line with the higher breakthrough infection risk observed in PLWH compared to HIV-negative controls (adjusted hazard ratio 1.28)[11]. In addition, a higher incidence of SARS-CoV-2 mortality was observed in PLWH (adjusted hazard ratio’s 3.29 and 2.59)[12, 13]. It was reported that the magnitude of the antibody response after vaccination correlates with protection against symptomatic infection with the ancestral viral strain[14]. This correlation likely holds true for novel emerging variants, although it is known that the neutralization potency diminishes with every new variant[15] and that two doses of BNT162b2 vaccination are not very effective in preventing symptomatic Omicron BA.1 or BA.2 infection[16]. With the continuing emergence of novel variants, it is reasonable to assume that antibodies remain important for clinical protection against SARS-CoV-2 infection. Indeed, in healthy individuals, one additional vaccination after a primary vaccination regimen restored neutralization potency against the subvariants BA.2.12.1 and BA.4/5[17]. Following from these observations and reasoning, PLWH may require additional vaccinations to achieve adequate protection against SARS-CoV-2 infection, especially with the emergence of novel antigenically distinct variants like the currently circulating Omicron lineage.

The effect of an additional SARS-CoV-2 vaccination on the humoral and cellular immune responses in PLWH with a low or absent serological response after completing a primary vaccination regimen is unknown. Three studies on additional SARS-CoV-2 vaccinations in PLWH have been performed that showed an increase in antibody levels, without analysis on the cellular immune response[18–20]. All three studies did not focus on those who may benefit most from additional vaccinations, namely the subgroup of hyporesponders after a primary vaccination regimen.

The aim of this study was to evaluate the SARS-CoV-2 spike (S)-specific immune responses after an additional mRNA-1273 vaccination in PLWH, who had a serological hyporesponse after a primary SARS-CoV-2 vaccination regimen.

## Methods

### Study design and participants

We conducted a nested single arm intervention trial embedded within the prospective nationwide cohort study in 22 of the 24 HIV treatment centres in the Netherlands (COVID-19 Vaccination response In people living with HIV, COVIH, n=1154). All 165 PLWH from the COVIH study with a hyporesponse (defined as ≤300 (S)-specific binding antibody units [BAU]/mL, measured at 4–6 weeks after a primary vaccination regimen with either two doses of BNT162b2, mRNA-1273, ChAdOx1-S, or one dose of Ad26.COV2.S) were eligible for participation. The serological assay cut-off definitions used here followed the consensus by the Dutch national expert working group (HARMONY) on the harmonization of SARS-CoV-2 immunological assays. Participants with evidence of intercurrent SARS-CoV-2 infection, demonstrated by a history of a reported or documented positive PCR or rapid antigen test, or with serological evidence of >300 BAU/mL shortly before the additional vaccination, were excluded.

### Clinical procedures

The intervention consisted of a single mRNA-1273 vaccination (100 µg) administered at the Erasmus University Medical Centre or Leiden University Medical Centre, both in the Netherlands. Blood samples were obtained immediately before the vaccination (at the same visit; T0) and 28 days later (T1) for collection of serum and peripheral blood mononuclear cells (PBMCs).

Clinical data were extracted from an electronic case record file. Recorded study variables included year of birth, sex assigned at birth, dates and type of primary SARS-CoV-2 vaccinations, current use of combination antiretroviral therapy (cART), most recent plasma HIV-RNA (copies/mL), most recent CD4+ T-cell count (cells/µL), nadir CD4+ T-cell count (cells/µL), and use of immunosuppressant medication.

Local and systemic adverse events were evaluated via a standardized printed or electronic diary concerning vaccination related side-effects and medication use in the seven days after the additional vaccination.

### Laboratory procedures

All serum samples were assessed at the Erasmus University Medical Centre (WHO SARS-CoV-2 reference laboratory) for the presence of SARS-CoV-2 S1-specific binding antibodies (hereafter S1-specific antibodies) with a validated IgG Trimeric chemiluminescence immunoassay (DiaSorin Liaison) with a lower limit of detection at 4.81 BAU/mL and a cut-off level for positivity at 33.8 BAU/mL.

SARS-CoV-2 S-specific neutralizing antibodies and S-specific T-cells were measured on a selection of 40 participants of whom 20 had their primary vaccination with ChAdOx1-S and 20 with BNT162b2. Selection was based on the S1-specific antibodies 28 days after the additional vaccination (T1), to represent the whole range of antibody responses as good as possible. Neutralizing functionality of antibodies was assessed by a plaque reduction neutralization test (PRNT) and the SARS-CoV-2 S-specific T-cells by an activation induced marker (AIM) assay (see Supplementary Appendix 1 for further details on laboratory tests).

SARS-CoV-2-S-fluorochrome-labelled tetramers were used to compare the S-specific B-cell compartment on a selection of 18 participants that were balanced for primary vaccination regimen (ChAdOx1-S or BNT162b2) and CD4+ T-cell counts. Four of these 18 participants were also included in a subgroup during the initial study, in which PBMCs were collected 21 days (+/- 3 days) after the first vaccination (AV1), and 4–6 weeks after completing a primary vaccination schedule (AV20). These PBMCs were used for longitudinal analysis of class-switching S-specific B-cells.

### Outcomes

The primary outcome was defined as the increase in S1-specific antibodies in PLWH 28 days after the additional vaccination compared to the S1-specific antibodies immediately prior to additional vaccination. An adequate response was defined as the presence of S1-specific antibodies >300 BAU/mL. Secondary outcomes included the association between patient characteristics and antibody responses, the detection of SARS-CoV-2 S-specific neutralizing antibodies targeting the ancestral SARS-CoV-2 (D614G) and Omicron (BA.1) variant, T-cell and B-cell responses targeting the ancestral SARS-CoV-2 (Wuhan-Hu1), and additionally T-cell responses targeting the Omicron (BA.1) variant. Lastly, we evaluated the tolerability by monitoring local and systemic vaccine related adverse events. Severity of reactogenicity was measured as mild (symptoms present but no functional impairment or medication needed), moderate (necessitating medication, no functional impairment) or severe (impairing daily functioning).

### Sample size and statistical analysis plan

The study was designed with the anticipation that 10% of the PLWH would have a serological hyporesponse after the primary vaccination series in the COVIH study. If we would be able to include 80 PLWH, we would have >95% power to detect a 20% increase in PLWH with an adequate serological response after the additional vaccination (one-sided alpha 0.05).

The baseline characteristics were described as number (percentage) or median (interquartile range [IQR]). The primary outcome was assessed as the difference between the S1-specific antibodies at T1 minus T0 with a 95% Confidence Interval (CI) by a Wilcoxon signed-rank test. We evaluated the proportion with adequate serological responses by a McNemar’s test. To investigate factors associated with the absolute increase in antibody response at 28 days after the additional vaccination in PLWH, we used unpaired *t* tests and a proportional odds generalized linear multivariable model with the covariates sex, age (subgroups 18-65 versus >65 years), most recent CD4+ T-cell count (subgroups <500 versus >500/mm^3^), nadir CD4+ T- cell count (subgroups <500 versus >500/mm^3^), and primary vaccination regimen (mRNA versus vector). 95% confidence intervals and p-values were reported for each coefficient in the regression model. Coefficients with p-values <0.05 were considered significant.

Undetectable serological responses (<4.81 BAU/mL) were reported as 4.81 in the statistical analyses, undetectable neutralizing antibodies (<10) as 10, and undetectable S-specific T-cell and B-cell responses (<0.01) as 0.01.

Data was analysed using GraphPad Prism 9.3.1. Flow cytometry data was analysed using FlowJo software version 10.8.1. FlowJo software was used to gate CD19+ B-cells and OMIQ data analysis software was used for further analysis (www.omiq.ai). Cytonorm was used for batch corrections followed by Uniform Manifold Approximation and Projection (UMAP) dimensionality reduction to visualize the phenotypes of S-specific B-cells.

## Results

### Baseline characteristics

Between 22^nd^ November 2021 and 28^th^ December 2021, 75 of the 165 invited PLWH were enrolled into this substudy. Before the additional vaccination was given, five (6.7%) participants had an increase in their S1-specific antibodies to >300 BAU/mL and were excluded. Two (2.7%) participants were excluded due to a PCR-confirmed SARS-CoV-2 infection within 28 days after the additional vaccination and two (2.7%) participants were lost to follow-up after having received the additional vaccination. Overall, 66 PLWH were included for the analysis. Of the 66 participants, 40 (60.6%) received ChAdOx1-S, 22 (33.3%) BNT162b2 and four (6.1%) Ad26.COV2.S as primary vaccination regimen (**Figure 1**).

**Figure 1.**
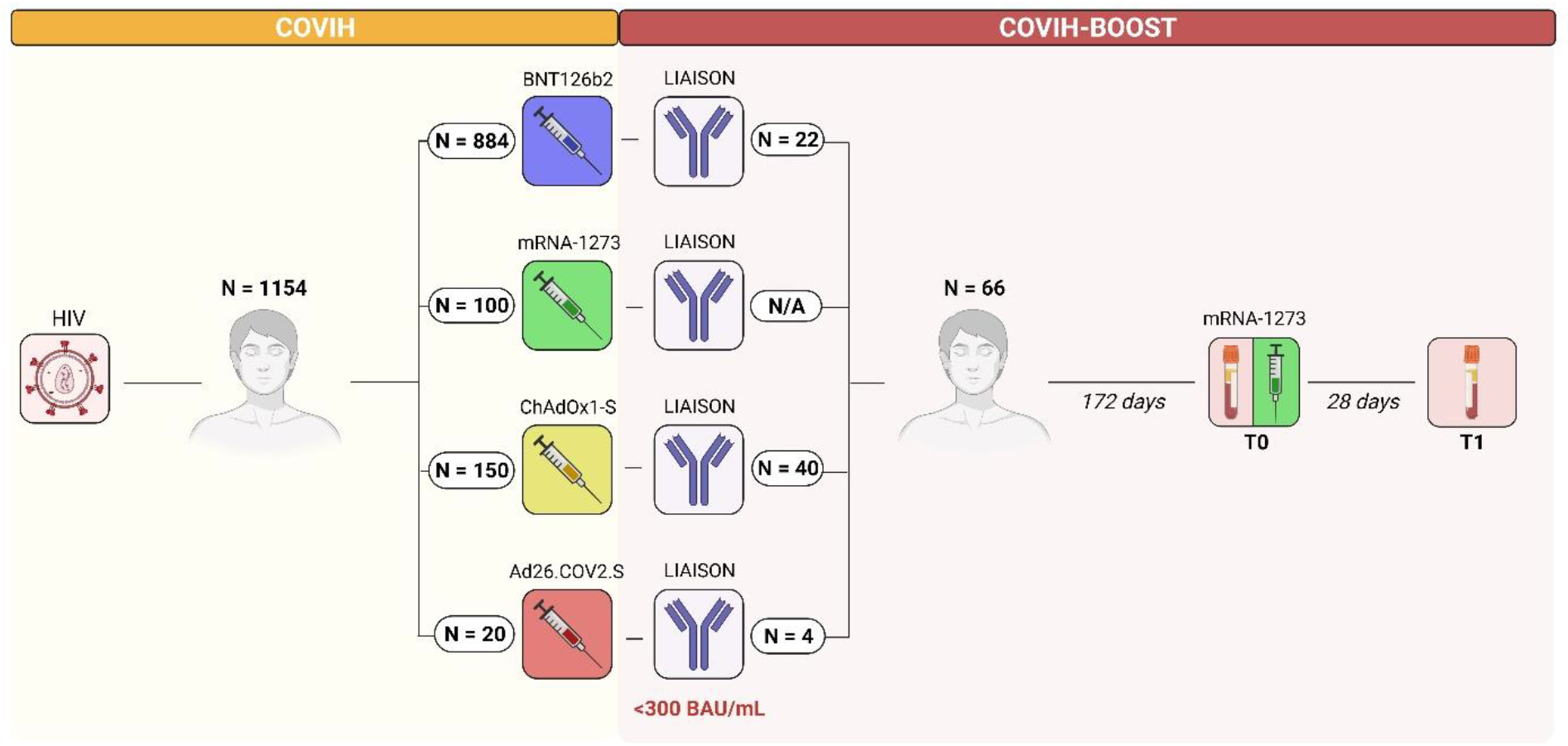
Study overview. Overview of the initial COVIH study and the COVIH-BOOST substudy with the number of included participants. A total of N=66 participants with hyporesponse after a primary vaccination regimen were enrolled in the substudy for additional mRNA-1273 vaccination and study of their immune response. N/A: not applicable.

Baseline characteristics of all participants are described in **Table 1**, including the characteristics of PLWH with hyporesponse from the initial study who were not enrolled in this substudy. Participants had a median age of 63 years [IQR:60-66], 86% were male and most recent median CD4+ T-cell count was 650 cells/μL [IQR:423-941] and nadir CD4+ T-cell count was 230 cells/μL [IQR:145-345]. The majority (97%) was on cART and had a suppressed plasma HIV-RNA (96% <50 copies/mL). No participants used immunosuppressant medication. Among the 66 participants analysed, the median time between completing the primary vaccination series and the additional vaccination was 172 days [IQR:154-195].

**Table 1.** Baseline characteristics of PLWH.

|  | **COVIH**<br>**participants** | **COVIH-BOOST participants** |  |  |
| --- | --- | --- | --- | --- |
|  | With<br>hyporesponse,<br>not included<br>N = 99 | With<br>hyporesponse,<br>included<br>N = 66 | Primary mRNA<br>vaccination<sup>a</sup><br>N = 22 | Primary vector<br>vaccination<sup>b</sup><br>N = 44 |
| **Sex assigned at birth** |  |  |  |  |
| Male | 87 (87.9) | 57 (86.4) | 20 (90.9) | 37 (84.1) |
| Female | 12 (12.1) | 9 (13.6) | 2 (9.1) | 7 (15.9) |
| **Age category** |  |  |  |  |
| 18-55 years | 33 (33.3) | 9 (13.6) | 5 (22.7) | 4 (9.1) |
| 56-65 years | 43 (43.4) | 34 (51.5) | 3 (13.6) | 31 (70.5) |
| 65+ years | 23 (23.2) | 23 (34.9) | 14 (63.6) | 9 (20.5) |
| **SARS-CoV-2 vaccinations in primary schedule** |  |  |  |  |
| BNT162b2 | 37 (37.4) | 22 (33.3) | 22 (100) | N/A |
| mRNA-1273 | 4 (4.0) | N/A | N/A | N/A |
| ChAdOx1-S | 42 (42.4) | 40 (60.6) | N/A | 40 (90.9) |
| Ad26.COV2.S | 16 (16.2) | 4 (6.1) | N/A | 4 (9.1) |
| **On cART** |  |  |  |  |
| Yes | 99 (100) | 64 (97.0) | 20 (90.9) | 44 (100) |
| **Most recent plasma HIV viral load** |  |  |  |  |
| <50 copies/mL | 93 (93.9) | 63 (95.5) | 20 (90.9) | 43 (97.7) |
| ≥50 copies/mL | 6 (6.1) | 3 (4.5) | 2 (9.1) | 1 (2.3) |
| **Most recent CD4+ T-cell count** |  |  |  |  |
| <250 cells/μL | 13 (13.1) | 5 (7.6) | 4 (18.2) | 1 (2.3) |
| 250-500 cells/μL | 21 (21.2) | 12 (18.2) | 5 (22.7) | 7 (15.9) |
| >500 cells/μL | 65 (65.7) | 49 (74.2) | 13 (59.1) | 36 (81.8) |
| **Nadir CD4+ T-cell count** |  |  |  |  |
| <250 cells/μL | 45 (45.5) | 31 (47.0) | 12 (54.5) | 19 (43.2) |
| 250-500 cells/μL | 32 (32.3) | 19 (28.8) | 5 (22.7) | 14 (31.8) |
| >500 cells/μL | 9 (9.1) | 7 (10.6) | 1 (4.5) | 6 (13.6) |
| Unknown | 13 (13.1) | 9 (13.6) | 4 (18.2) | 5 (11.4) |
| **Use of immunosuppressant medication** |  |  |  |  |
| Yes | 4 (4.0) | 0 | 0 | 0 |
| **Days since completing primary vaccination** | N/A | 172 [154-195] | 192 [165-206] | 168 [152-181] |

Values are number (percentage) for categorical variables or median [IQR] for continuous variables. <sup>a</sup>BNT162b2, mRNA-1273. <sup>b</sup>ChadOx1-S, Ad26.COV2.S. N/A: not applicable, cART: combination antiretroviral therapy, IQR: interquartile range.

The mean S1-specific antibody level directly pre-vaccination was 35 BAU/mL (SEM 5.4), including 45 participants with serological responses below the cut-off of the test (<33.8 BAU/mL). Following ChAdOx1-S primary vaccination the antibody level directly pre-vaccination was mean 31 BAU/mL (SEM 5.4), 46 BAU/mL (SEM 12.8) after BNT162b2 and 19 BAU/mL (SEM 6.3) after Ad26.COV2.S.

### S1-specific antibodies

Twenty-eight days [IQR:28.0-28.0] after the additional vaccination, S1-specific antibodies >300 BAU/mL were measured in 64/66 (97.0%) of the participants (p<0.0001). All participants showed an increase in S1-specific antibodies after vaccination, which was mean 4282 BAU/mL (95%CI:3241-5323, p<0.0001) (**Figure 2**).

**Figure 2.**
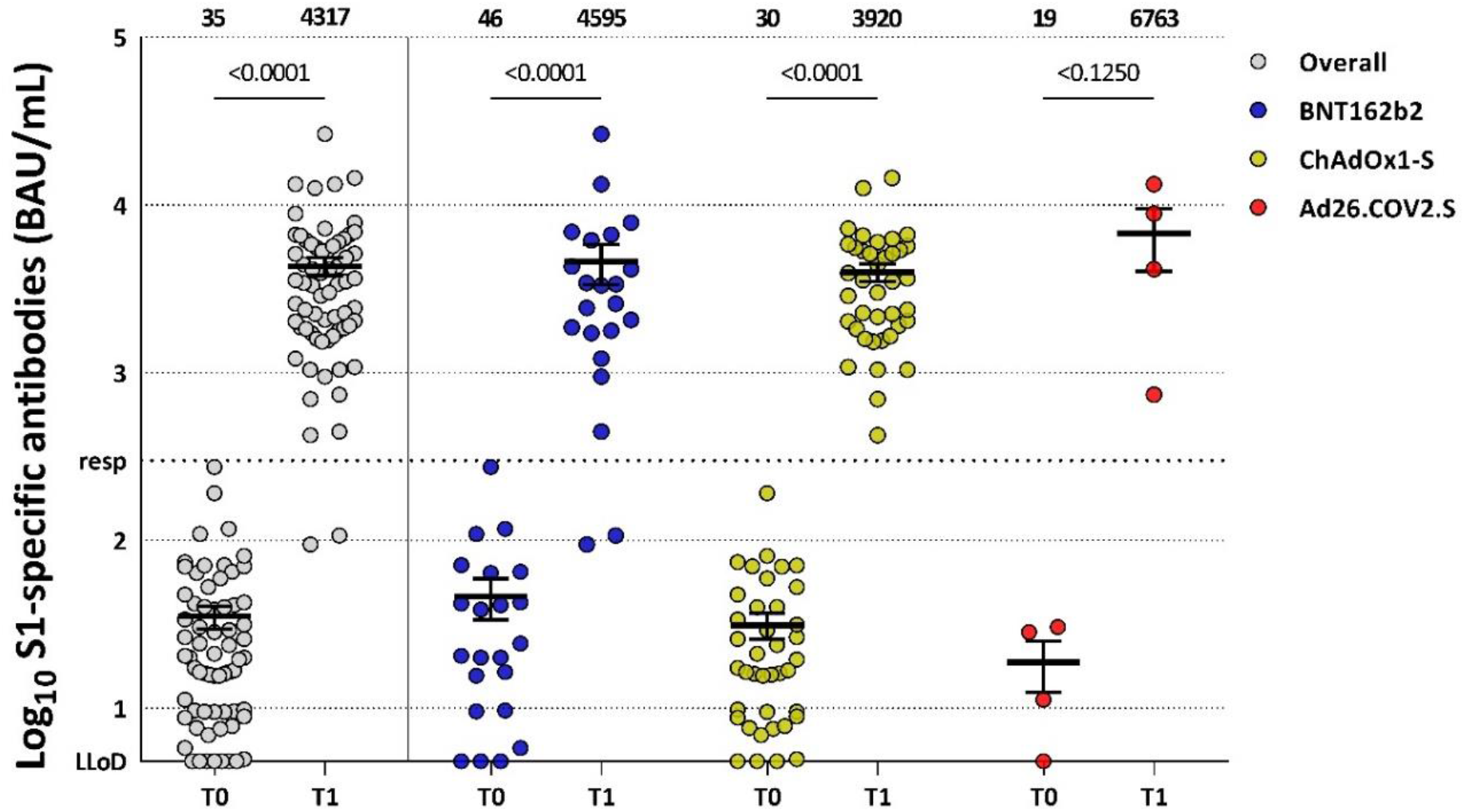
SARS-CoV-2 S1-specific binding antibody levels in PLWH after additional mRNA-1273 vaccination. Levels of S1-specific binding antibodies measured 28 days after the additional mRNA-1273 vaccination in all 66 PLWH (grey), in 22 PLWH after primary vaccination with BNT162b2 (blue), in 40 PLWH after primary vaccination with ChadOx1-S (yellow), and in four PLWH after primary vaccination with Ad26.COV2.S (red). The thick horizontal bar shows the mean S1-specific binding antibody level, also indicated above the graph, with error bars showing the standard error of the mean. LLoD is 4.81 BAU/mL, adequate responder (resp) cut-off is 300 BAU/mL (dotted line). Comparisons of timepoints were performed by paired *t* test. S: spike, LLoD: lower limit of detection, T0: before additional vaccination, T1: 28 days after additional vaccination.

Only two participants with a well suppressed HIV on cART did not reach >300 BAU/mL after the additional vaccination with S1-specific antibodies increasing from 8.71 and <4.81 BAU/mL to 94.7 and 107 BAU/mL, respectively. Both participants received BNT162b2 as primary vaccination and were males with low CD4+ T-cell nadirs <50 cells/µL and most recent CD4+ T-cell counts of 230 cells/µL and 313 cells/µL.

The mean increase in S1-specific antibodies was comparable between primary vaccination with ChAdOx1-S, BNT162b2 or Ad26.COV2.S. A mean increase of 3890 BAU/mL (95%CI:2945-4835) was measured after primary vaccination with ChAdOx1-S, 4549 BAU/mL (95%CI:1986-7112) after primary vaccination with BNT162b2 and 6744 BAU/mL (95%CI:-1978-15465) after primary vaccination with ad26.COV2.S (p=0.57). The mean increase in S1-specific antibodies was also not significantly different between most recent CD4+ T-cell count of <500 versus >500/mm^3^ (5400 BAU/mL versus 3895 BAU/mL, p=0.85), age 18-65 versus >65 years (4336 BAU/mL versus 4183 BAU/mL, p=0.52) and male versus female (4372 BAU/mL versus 3714 BAU/mL, p=0.42).

A proportional odds generalized linear regression model was performed to investigate factors associated with the absolute increase in antibody response 28 days after the additional vaccination. This adjusted analysis did not identify significant associations between any of the patient characteristics of interest and S1-specific antibody responses (**Table 2**).

**Table 2.** Multiple linear regression model of the association between antibody response and different variables in PLWH (N=66) after additional vaccination. The association between patient-related and vaccine-related variables on the absolute increase in S1-specific binding antibodies 28 days after the additional mRNA-1273 vaccination in PLWH with hyporesponse after a primary vaccination regimen was investigated by means of a proportional odds generalized linear regression model.

**S1-specific binding antibodies 28 days after additional vaccination in 66 PLWH**
| Variable | Estimate | 95% CI | p-value |
| --- | --- | --- | --- |
| Male sex | 869.1 | -2483 to 4221 | 0.61 |
| Age >65 vs 18-65 years | -576.3 | -3851 to 2698 | 0.73 |
| Most recent CD4+ T-cell count<br><500/mm<sup>3</sup> vs >500/mm<sup>3</sup> | 1301 | -1257 to 3859 | 0.31 |
| Nadir CD4+ T-cell count <500/mm<sup>3</sup><br>vs >500/mm<sup>3</sup> | -1175 | -4288 to 1938 | 0.45 |
| Primary vaccination type mRNA | -523.4 | -3498 to 2452 | 0.73 |

### Neutralizing antibodies

After additional vaccination, neutralizing antibodies against the ancestral SARS-CoV-2 were present in all subgroup participants (40/40) and against the Omicron (BA.1) variant in 65% of participants (26/40) (**Figure 3A**). Neutralizing antibodies against the ancestral virus were higher when participants had primary vaccination with ChadOx1-S compared to primary vaccination with BNT162b2, mean PRNT50 of 3526 versus 1611 (p=0.003). Neutralizing antibodies against the circulating Omicron variant were numerically higher as well after ChadOx1-S, mean PRNT50 of 889 versus 442 (p=0.46).

**Figure 3.**
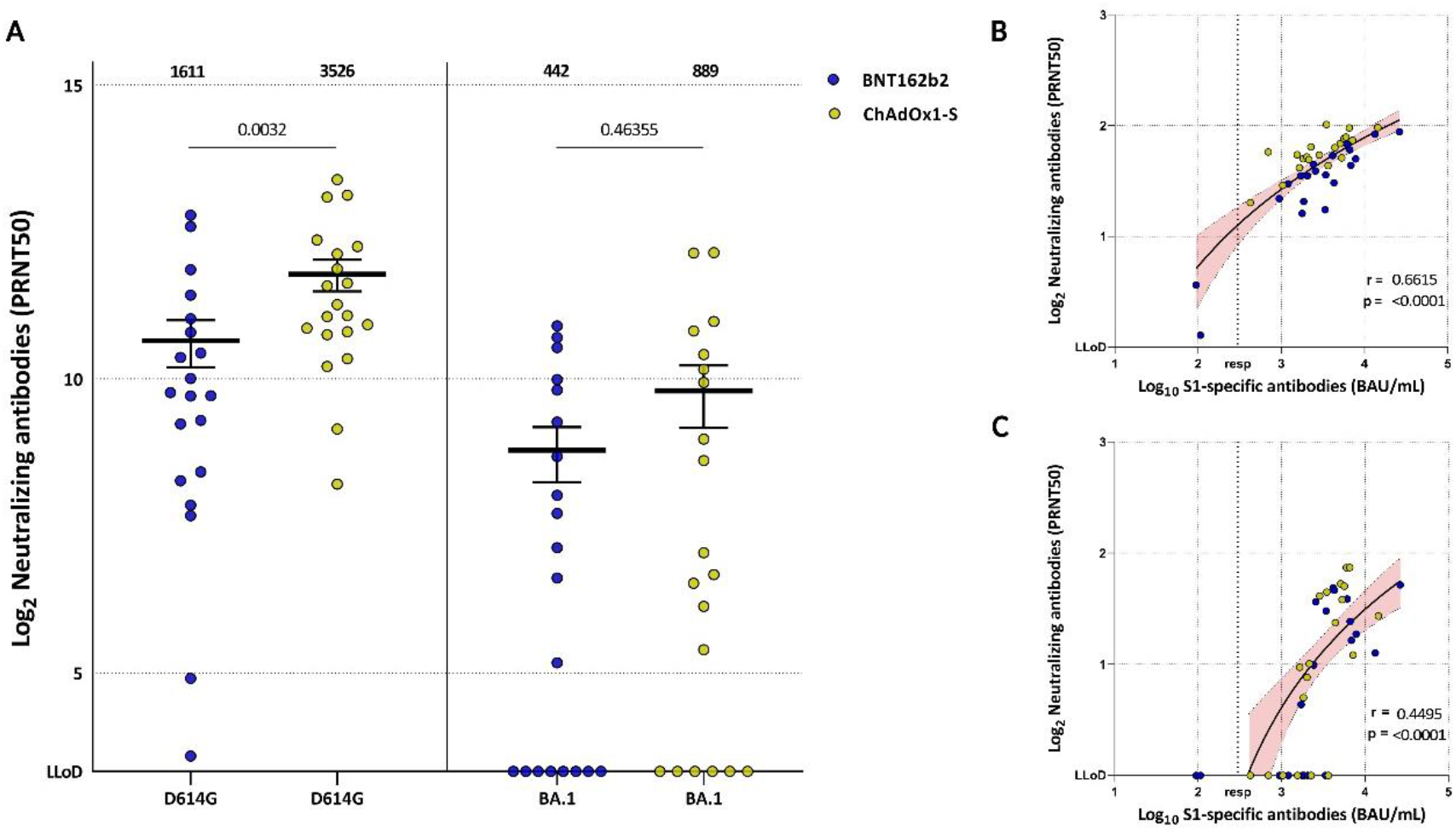
Neutralizing antibodies to SARS-CoV-2 in subgroup participants (N=40) after additional mRNA-1273 vaccination. (A) PRNT50 titer measured 28 days after the additional mRNA-1273 vaccination against the ancestral SARS-CoV-2 (D614G) and Omicron (BA.1) variant after primary vaccination with ChAdOx1-S (yellow) and BNT162b2 (blue). The thick horizontal bar shows the mean neutralizing antibody titer, also indicated above the graph, with error bars showing the standard error of the mean. LLoD is 10. Comparisons between the two different primary vaccination groups were performed using unpaired *t* test. (B) Correlation between the S1-specific binding antibody levels and neutralizing antibody levels targeting the ancestral SARS-CoV-2 by a linear regression analysis on transformed data, R=0.66, p<0.0001. (C) Correlation between the S1-specific binding antibody levels and neutralizing antibody levels targeting the Omicron BA.1 variant by a linear regression analysis on transformed data, R=0.45, p<0.0001. Adequate responder (resp) cut-off is 300 BAU/mL (dotted line). LLoD: lower limit of detection, S: spike.

A positive correlation between S1-specific antibodies and the neutralizing antibodies against the ancestral variant (R = 0.66, p<0.0001) was observed (**Figure 3B**). That correlation was less clear for neutralizing antibodies against the Omicron variant (R = 0.45, p<0.0001) (**Figure 3C**).

### Frequency of SARS-CoV-2 S-specific T-cells

Before additional vaccination high frequencies of ancestral S-specific T-cells (median 0.075% [IQR:0.010-0.210]) and Omicron S-specific T-cells (median 0.055% [IQR:0.013-0.235]) were detected. Additional vaccination led to a non-significant increase in ancestral S-specific CD4+ T-cells (median 0.075%, p=0.51) (**Figure 4**). However, the proportion of PLWH with detectable ancestral S-specific CD4+ T-cells significantly increased from 27/40 (67.5%) before additional vaccination (T0) to 34/39 (87.2%) after additional vaccination (T1) (p=0.037). Omicron S-specific CD4+ T-cells were comparable before and after additional vaccination (p=0.95), and also the proportion of PLWH with detectable Omicron S-specific CD4+ T-cells did not differ between T0 and T1 (p=0.945).

**Figure 4.**
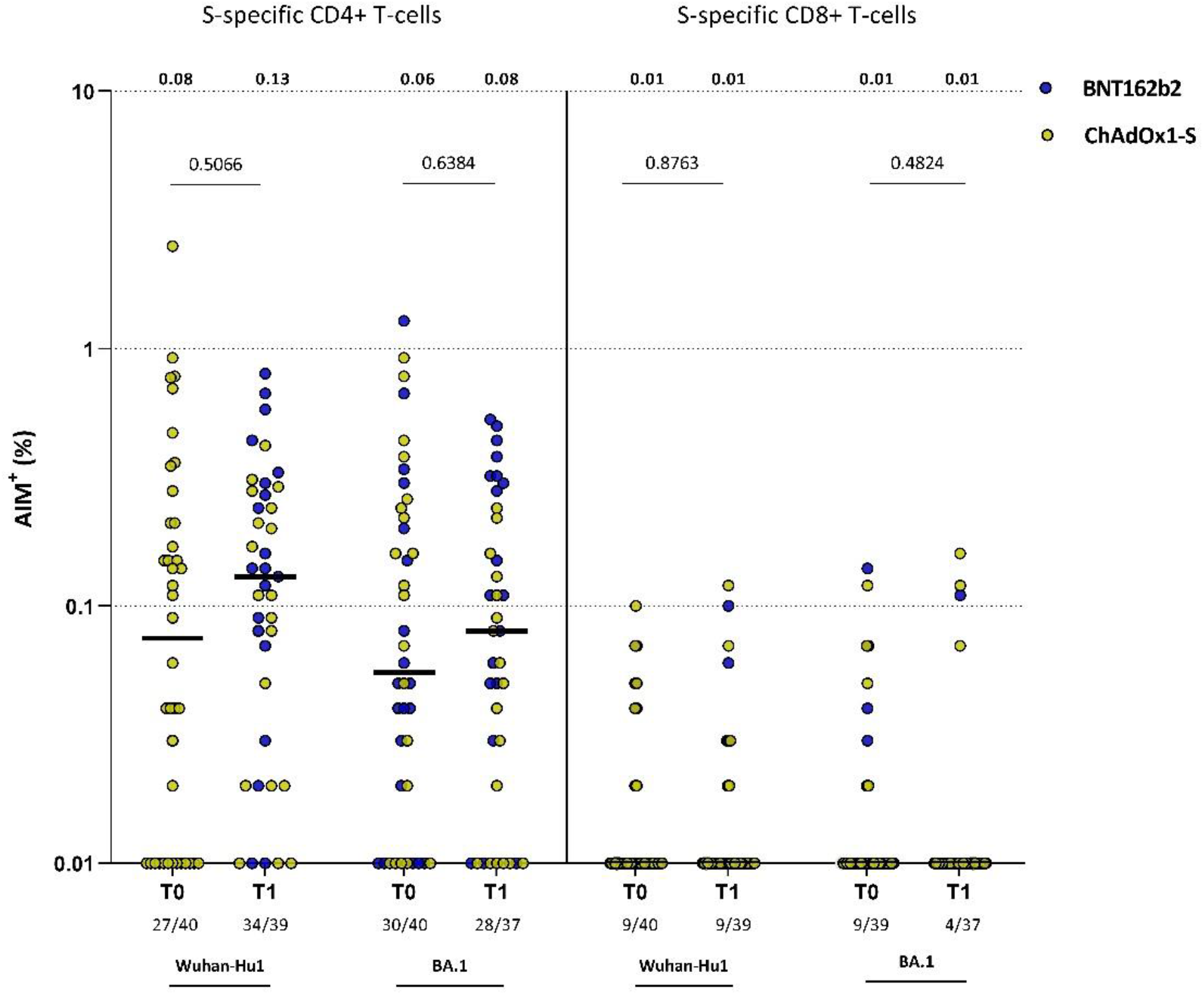
Frequency of SARS-CoV-2 S-specific T-cells in subgroup participants (n=40) before and 28 days after additional mRNA-1273 vaccination. CD4+ (CD4+CD40L+CD137+) and CD8+ (CD8+CD69+CD137+) T-cell responses to ancestral spike (Wuhan-Hu1) and Omicron spike (BA.1) measured by the AIM assay before additional vaccination (T0) compared to 28 days after additional vaccination (T1). Participants who received ChAdOx1-S as primary vaccination are shown in yellow, participants who received BNT162b2 as primary vaccination are shown in blue. The horizontal line shows the median, also indicated above the graph. The total numbers of participants with detectable S-specific T-cells are indicated below the graphs. Comparisons of timepoints were performed by unpaired *t* test. AIM: activation induced marker assay, S: spike, T0: before additional vaccination, T1: 28 days after additional vaccination.

S-specific CD8+ T-cells were infrequently observed in study participants and no effect on S-specific CD8+ T-cells was observed after additional vaccination for both the ancestral SARS-CoV-2 (p=0.876) and the Omicron variant (p=0.482).

### Frequency and phenotype of SARS-CoV-2 S-specific B-cells

Low frequencies (median 0.017% [IQR:0.001-0.034]) of S-specific B-cells were detected at T0. S-specific B-cells significantly increased (median 0.258% [IQR:0.126-0.488], p=0.0003) 28 days after participants received the additional vaccination (**Figure 5A**). The proportion of PLWH with detectable S-specific B-cells also significantly increased from 13/18 (72.2%) at T0 to all participants (100%) at T1 (p=0.016). To further investigate the phenotype of S-specific B-cells, we compared the frequencies of unswitched (IgD, IgM and IgMD) and switched (IgG and IgA) S-specific B-cells (Supplementary Figure 3). Although not many S-specific B-cells were present before the additional vaccination, the majority was already class-switched to IgG (mean 61%). However, a significant increase in switched IgG (mean 82%, p=0.013) was observed 28 days after the additional vaccination (**Figure 5B**). Longitudinal analysis of class-switching of S-specific B-cells suggests a decrease in switched IgG+ S-specific B-cells after a primary vaccination regimen in the PLWH with initial hyporesponse (Supplementary Figure 4). Because the majority of S-specific B-cells class-switched to IgG, we further focused on this memory IgG-compartment. As expected, percentages of IgG+ S-specific B-cells correlated with the S1-specific antibodies (**Figure 5C**). However, percentages of IgG+ S-specific memory B-cells did not correlate with the most recent CD4+ T-cell count (Supplementary Figure 5). To investigate the phenotypical properties of IgG+ S-specific B-cells (e.g. migratory capacity (CXCR3), proliferating (Ki67), activation status (CD95), recent germinal center graduation (CD21-)[21], and memory subsets, like CD45RB+ B-cells and CD11c+CD19^high^ cells[22]), we used a panel of 24 antibodies. In total, 6004 S-specific B-cells were measured from all samples. To visualize how the phenotype of S-specific B-cells changed after additional vaccination, we performed Uniform Manifold Approximation and Projection (UMAP) dimensionality reduction and depicted the S-specific B-cells per timepoint in black (**Figure 5D**). Activated (CD95+), Migrating (CXCR3+) and memory (CD11c+CD19^high^) S-specific IgG+ B-cells were significantly most prevalent at T1, as were B-cells recently graduating from the germinal centers (CD21-B-cells). Proliferating (Ki67) S-specific IgG+ B-cells were not significantly different between T0 and T1 (**Figure 5E**, Supplementary Figure 6).

**Figure 5.**
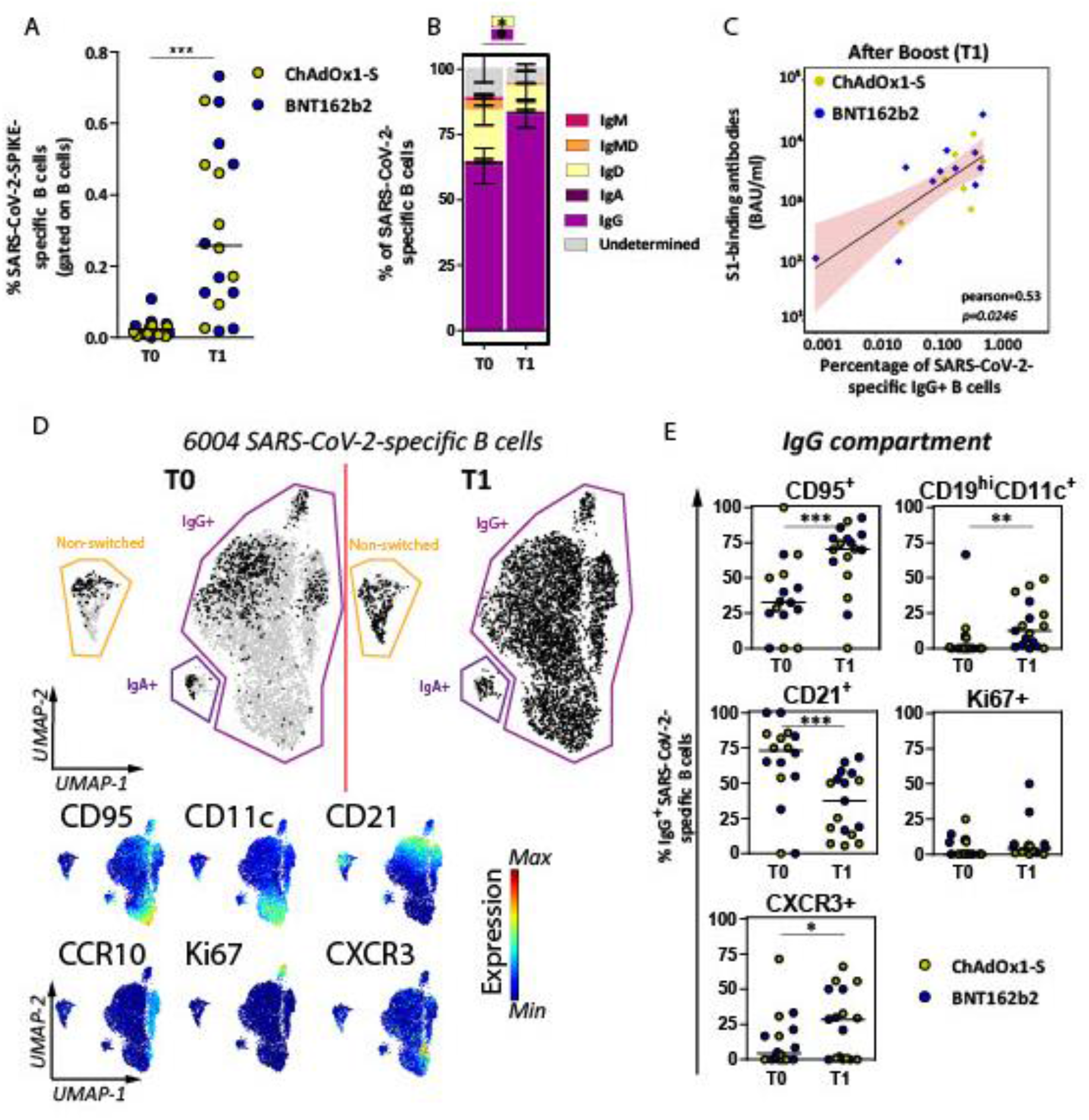
Frequency and phenotype of SARS-CoV-2 S-specific B-cells in subgroup participants (n=18) before and 28 days after an additional mRNA-1273 vaccination. (A) Percentages of S-specific B-cells are shown as frequencies from total B-cells per individual and each individual is colored according to the primary vaccination regimen (yellow: ChAdOx1-s (n=8) and blue: BNT162b2 (n=10)). The horizontal line shows the median. (B) Isotype usage of S-specific B-cells are shown as stacked bars at T0 and T1. (C) Correlation plot between S1-specific binding antibody levels and IgG+ S-specific B-cells. Pearson correlation analysis on non-transformed data is depicted and linear regression results shown as a black line with red shaded 95% confidence intervals. (D) Uniform Manifold Approximation and Projection (UMAP) for all 6004 S-specific B-cells (grey) to cluster cells based on 24 different markers. S-specific B-cells are overlaid based on timepoint (black) on top of all cells (grey). Normalized expression of six selected markers is shown below the overlaid UMAP. Expression plots for all markers can be found in Supplementary Figure 6. (E) Manually gated B-cell subsets are shown within the IgG+ B-cell compartment at each timepoint. Statistical analyses, Wilcoxon matched pairs tests were performed for panels A, B and E to compare T0 vs T1. *=p<0.05, **=p<0.01, ***=p<0.001. Individuals and median values are shown for panels A and E. Means with standard error of means are shown in panel B.

### Reactogenicity

The additional dose was well tolerated and no serious adverse events were reported. Overall, 66.1% of the participants reported local or systemic adverse events, with pain at the injection site as most frequent local reaction and generalised myalgia and headache as most frequent systemic reactions (Supplementary Figure 3).

## Discussion

To our knowledge, this study is the first to report the immunogenicity of an additional SARS-CoV-2 vaccination in PLWH who had a low antibody response after a primary vaccination regimen. A substantial increase in S1-specific antibodies and memory B-cells was shown, supporting the usefulness of additional booster vaccinations in PLWH and in particular for those with a documented hyporesponse after a primary vaccination regimen.

Remarkably, there was no significant association between the increase in S1-specific antibodies and the primary vaccination regimen, most recent CD4+ T-cell count, nadir CD4+ T-cell count, age, and sex. However, smaller differences between these variables cannot be ruled out since the study was powered on overall responsiveness and not for absolute differences in S1-specific antibodies between groups.

As expected, a positive correlation was observed between neutralization potency and S1-specific antibodies. This supports the observation that the correlation between the magnitude of the antibody levels and neutralization for the ancestral strain is also true in PLWH for variants. Since neutralization was reported to be a correlate of protection against infection and disease, our data support a strategy to obtain an ultimate level of antibodies for optimal protection against SARS-CoV-2 infection in PLWH.

S-specific T-cells were detected before the additional vaccination in the majority of PLWH, despite the low levels of S1-specific antibodies. This implicates that although it is important to boost S1-specific antibodies in hyporesponders, there still is a second line of defence. After an additional vaccination, the proportion of participants with detected CD4+ T-cells targeting the ancestral variant significantly increased, but not for the Omicron variant and neither was such effect observed for CD8+ T-cell responses. However, it is known that these S1-specific T-cells against ancestral SARS-CoV-2 cross-react with Omicron[23].

Furthermore, there was a significant induction of activated/homing and memory S-specific B-cells after additional vaccination that correlated with the S1-specific antibodies. Combined with the longitudinal B-cell data from four participants, this implicates that an additional vaccination is needed in these PLWH with hyporesponse to induce activation and memory B-cells for durable protection.

Other studies of additional SARS-CoV-2 vaccinations in different immunocompromised patient groups with a hyporesponse after a primary vaccination regimen also show that additional vaccination led to an increase in antibody levels. Nonetheless, lower seroconversion rates above a predefined cut-off were observed[24, 25]. However, the cut-offs for response differed between the studies and some studies only included participants with an antibody level <50 BAU/mL after a primary vaccination regimen, making the results less comparable with this study. A comparable study in patients receiving chemotherapy, immunotherapy, or both for solid tumours showed that 46 of the 48 hyporesponders reached S1-specific antibodies >300 BAU/mL 28 days after a third mRNA-1273 vaccination[26]. Therefore, compared to the literature on additional vaccinations in immunocompromised patients’ groups, PLWH that are on an effective cART regimen with hyporesponse after a primary vaccination regimen appear to seroconvert more frequently and with higher increases in magnitude of S1-specific antibody response after an additional vaccine dose.

Our study had several limitations. First, our study population had an imbalance in sex distribution. Participants were generally virally suppressed on cART with CD4+ T-cell counts above 500 cells/μL, limiting the generalisability to the overall population of PLWH. Furthermore, the absence of a control group should be noted. Measuring the S1-specific antibodies is no standard policy in the healthy population, making it difficult to compose a control group of hyporesponders. Although we did not meet our predefined sample size for a study power of >95%, the observed effect size was considerably higher than anticipated. As a consequence, the risk of an underpowered study is very unlikely. Our results show the immune responses before and 28 days after an additional vaccination. Further follow-up will take place at 6, 12, 18 and 24 months after the additional vaccination to determine the waning of immune responses in PLWH, which is important as faster waning of some other vaccines has been observed in this population[27]. Decay of S1-specific antibodies and T-cell responses has been described in a small study population of eight PLWH compared to 25 controls two weeks and six months after a primary vaccination regimen[28]. Virally suppressed PLWH with high CD4+ T-cell counts showed persistent S1-specific T-cell responses and a significant decline in S1-specific antibodies, both similar to healthy controls. However, our follow-up will take place in a larger study population, with both initial hyporesponders and adequate responders, and with different vaccination regimens. Ideally, our results should be confirmed by clinical outcome studies in PLWH. Therefore, we will also collect clinical data during the two-year follow-up, including the incidence of breakthrough infections.

In conclusion, an additional mRNA-1273 vaccination substantially improved humoral and cellular immune response in PLWH with low SARS-CoV-2 antibodies after a primary vaccination regimen. This shows that additional vaccinations are an effective approach in compensating for the reduced antibody responses in PLWH. In addition, the results of this study indicate that also primary vaccine responsive people with waning vaccine induced responses can expect benefit from future boosters to reinforce protection against infection with viral variants.

## Data Availability

All data produced in the present study are available upon reasonable request to the authors.

## Footnotes

### Conflict of interest statements outside the submitted work

SJ received grants from the Dutch research council (NWO), EU (Horizon 2020) and the Bill and Melinda Gates foundation. WFWB declares reimbursement for participation of patient in trial by GSK to institution. DG and RDdV are supported by the Health∼Holland grant EMCLHS20017 co-funded by the PPP Allowance made available by the Health∼Holland, Top Sector Life Sciences & Health, to stimulate public–private partnerships. RDdV is listed as inventor of the fusion inhibitory lipopeptide [SARS_HRC_-PEG_4_]_2_-chol on a provisional patent application. KCES received honorariums for advisory boards from Gilead and ViiV. CR has received research grants from ViiV, Gilead, ZonMW, AIDSfonds, Erasmus MC, and Health∼Holland and honorariums for advisory boards from Gilead and ViiV. BJAR declares research grants from Gilead and MSD and honorary for advisory boards for AstraZeneca, Roche, Gilead, F2G. KB received research and educational grants from ViiV, Gilead, and consulting fees for advisory boards for ViiV, Gilead, MSD and AstraZeneca. AR received grants from the Bill and Melinda Gates foundation and the Leids Universitair Fonds, participated on a board of investor-initiated clinical trial on convalescent plasma for COVID-19 and is chief editor of the Dutch journal of Infectious Diseases and member of the EMA expert group vaccines.

All other authors declare no conflicts of interest.

### Ethics committee approval

The trial was performed in accordance with the principles of the Declaration of Helsinki, Good Clinical Practice guidelines, and in accordance with the Dutch Medical Research Involving Human Subjects Act (WMO). Participants signed an extra informed consent form for participation in the substudy. The trial was reviewed and approved by the Medical research Ethics Committees United Nieuwegein (MEC-U, reference 20.125). The trial was registered in the Netherlands Trial Register (NL9214).

### Data sharing statement

Individual participant data that underlie the results reported in this article, after de-identification, will be made available to researchers who provide a methodologically sound study proposal.

### Authors’ contributions

Conceptualization: MJJ, KSH, DG, BJAR, KB, CR, AHER, YMM, PDK, RDdV, CHGvK

Formal analysis: MJJ, DG, WH, GP

Funding acquisition: BJAR, KB, CR, AHER, RDdV

Investigation: MJJ, KSH, DG, WH, SB, LG, JGdH, EFS, HSMA, WFWB, MvdV, MAHB, RS, NL, AHWB, EML, KCES, MGAvV, CED, JB, BJAR, KB, CR, AHER

Methodology: MJJ, KSH, BJAR, KB, CR, AHER, YMM, PDK, GP, RDdV, CHGvK

Project administration: MJJ, KSH, BJAR, KB, CR, AHER

Supervision: BJAR, KB, CR, AHER, YMM, RDdV, CHGvK

Validation: MJJ, KSH, DG, CR, BJAR, KB, CR, AHER, YMM, RDdV, CHGvK

Visualisation: MJJ, DG, KSH, WH, BJAR, KB, CR, AHER

Writing – original draft: MJJ, KSH, WH, BJAR, KB, CR, AHER

Writing – review & editing: all authors contributed to reviewing and editing of the manuscript

## Acknowledgements

Foremost, we would like to thank all participants of the study. We also want to thank the following people for recruitment of participants: Femke van Malsen, Roos van Heerde, Natasja van Holten, Willemien Dorama, Maartje Wagemaker, Ayten Karisli, René van Engen, Vincent Peters, Suzanne de Munnik, Vera Maas, Laura Laan, Jasmijn Steiner, Leontine van der Prijt and Jolanda van der Swaluw. Finally, we would like to thank Alessandro Sette and Alba Grifoni (La Jolla Institute for Immunology, La Jolla, San Diego, USA) for providing the peptide pools used in the AIM assay.

