## Supplemental Materials for "Immunogenicity of an additional mRNA-1273 SARS-CoV-2 vaccination in people living with HIV with hyporesponse after primary vaccination"

Supplementary Materials

### Supplementary Methods

#### Supplementary Appendix 1. Laboratory procedures

##### Plaque reduction neutralization test (PRNT)

Heat-inactivated serum was 2-fold diluted starting at 1:10 in OptiMEM medium supplemented with Glutamax, penicillin (100 IU/mL) and streptomycin (100 IU/mL). 400 plaque forming units SARS-CoV-2 ancestral (D614G) or Omicron (BA.1) were added to diluted sera and incubated at 37°C for one hour. Following incubation, virus-antibody mixture was transferred onto Calu-3 cells and incubated at 37°C for another eight hours. Next, cells were fixed in 4% paraformaldehyde (PFA; Avantor) and plaques were stained using a polyclonal rabbit anti-SARS-CoV-2 nucleocapsid antibody (Sino Biological) and a secondary peroxidase-labeled goat anti-rabbit IgG (Dako). Signal was developed by using a precipitate-forming 3,3′,5,5′-tetramethylbenzidine substrate (TrueBlue; Kirkegaard & Perry Laboratories) and the number of infected cells was counted per well by using an ImmunoSpot Image Analyzer (CTL Europe GmbH).

##### PBMC isolation

PBMCs were isolated by density gradient centrifugation (Ficoll-Hypaque, GE Healthcare life sciences) and collected in RPMI-1640 (Life Technologies) supplemented with 3% foetal bovine serum (FBS). PBMCs were washed three times before counting. Cells were frozen in freezing media (90% FBS supplemented with 10% dimethyl sulfoxide (DMSO)) and stored in liquid nitrogen until use.

##### SARS-CoV-2 S-specific T-cell analysis by activation induced marker (AIM) assay

PBMCs were thawed in RPMI-1640 medium supplemented with 10% FBS, penicillin (100 IU/mL), streptomycin (100 IU/mL), and L-glutamine and incubated with Benzonase® (50 IU/mL; Merck) at 37°C for 30 minutes. 1x10^6^ PBMCs were incubated with peptide pools (1µg/mL per peptide) and incubated at 37°C for 20 hours. Peptide pools consisted of 15-mers with 10 amino acid overlap covering the S-protein of the WuhanHu1 (ancestral) or B.1.1.529 (Omicron BA.1) variant and were a kind gift from Alessandro Sette and Alba Grifoni, La Jolla institute for Immunology. PBMCs were stimulated with an equimolar amount of DMSO as negative control or a combination of PMA (50 µg/mL) and Ionomycin (500 µg/mL) as positive control. Additionally, a CEFX Ultra SuperStim peptide pool (JPT) was included as a positive control. The CEFX peptide pool consisted of 176 peptides from common human pathogens and commensals. Following stimulation, cells were stained for phenotypic lymphocyte markers, memory markers, and activation markers and were measured by flow cytometry (FACSLyric, BD). PBMCs were stained for surface markers at 4°C for 15 minutes with 50µl of extracellular antibody cocktail (Supplementary Table 1). T-cells were gated as LIVE CD3+ cells and subdivided into CD4+ or CD8+ cells. CD45RA+CCR7+ (naïve, TN) were excluded from analysis and S-specific T-cells were identified as CD137+Ox40+ for CD4+ or CD137+CD69+ for CD8+ within the memory T-cell population [CD45RA-CCR7+ (central memory, TCM), CD45RA-CCR7- (effector memory, TEM), or CD45RA+CCR7- (terminally differentiated, TEMRA)]. The gating was set based on the DMSO stimulated sample on a per donor basis (Supplementary Figure 1).

##### Generation of SARS-CoV-2 S-labelled tetramers

Fluorescently labelled tetramers were generated by labelling biotinylated SARS-CoV-2-S protein (R&D, cat#10549-050) with streptavidin-coupled BUV615 (BD, cat#613013) or streptavidin-coupled BUV661 (BD, cat#612979). SARS-CoV-2-S protein was resuspended in 100µl PBS to get a 500µg/ml stock mixture (3730pmol/ml). SARS-CoV-2-S protein was then diluted two times to get a 1865pmol/ml user solution. Biotinylated SARS-CoV-2-S protein was then mixed in an approximate 4:1 molecular ratio with each of the two streptavidin-coupled fluorochromes (at 100µg/ml). Incubation was performed on ice in a stepwise approach where one-tenth fraction of streptavidin-coupled fluorochrome was added to the biotinylated SARS-CoV-2-S protein every 10 minutes. At 50 and 100 minutes the tetramers were spun down using a short pulse-spin. After the final incubation, free biotin was added to a final concentration of 30µM and incubated for 30 minutes on ice to block all unbound streptavidin-coupled fluorochromes. Labelled SARS-CoV-2-S tetramers (91µg/ml) were stored in the dark at 4°C and used within two weeks.

##### SARS-CoV-2 S-specific B-cell analysis

For immunophenotyping of S-specific B-cells, 3-5x10^6^ PBMCs were thawed in pre-warmed RPMI (Gibco) containing 20% heat-inactivated Fetal Bovin Serum (FBS; Lonza ) and washed once in the same media. All centrifugation steps of non-fixed samples were performed at 450g. Cells were transferred to a 96wells well, washed with PBS and then resuspended in 100µl PBS containing 1/500 diluted LIVE/DEAD fixable Blue Dead Cell Stain (Invitrogen Cat#L23105) and 1/50 diluted Fc Receptor binding inhibitor (eBioscience cat# 14-9161-73) for 15 minutes at roomtemperature in the dark. Then, samples were washed with PBS containing 2mM EDTA and 0.5% BSA followed by resuspension in 100µl of extracellular antibody cocktail (Supplementary Table 2) for 15 minutes at roomtemperature in the dark. This cocktail includes streptavidin-coupled BV421 (BD, cat#563259) as a ‘decoy probe’ to remove B-cells that react with streptavidin. Then, stained samples were washed and resuspended into 100µl of 2ug/ml of BUV615-labeled and BUV661-labeled S-specific tetramers for 30 minutes on ice in the dark. For the intracellular antibody staining, cells were first resuspended in FoxP3 fixation/permeabilization solution (Invitrogen, cat#00-5521-00) and incubated for 30 minutes on ice in the dark, followed by three washes with once permeabilization Buffer (Invitrogen, cat#00-8333-56). All centriguation steps of fixed samples were performed at 800g. Then, samples were resuspended in intracellular antibody cocktail, containing Ki67-BV711 (Biolegend, cat#350515), IRF4-APC (Miltenyi, cat#130-100-915) and caspase-3-V450 (BD, cat#560627). Fixed cells were washed with PBS containing 2mM EDTA and 0.5% BSA and 1-3x10^6^ events were acquired on a Cytek Aurora 5L spectral flow cytometer and unmixed using SpectroFlo software. Two samples were excluded due to the low number of events. See Supplementary Table 2 for details on the type of single-stain reference controls used for unmixing. For gating strategies for SARS-CoV-2 S-specific B cells see Supplementary Figure 2.

### Supplementary Tables

#### Supplementary Table 1. Panel of different monoclonal antibodies used for phenotypical analysis of SARS-CoV-2 S-specific T-cells.

|  | **Marker** | **Fluorochrome** | **Clone** | **Company** | **Catalogue** | **Dilution** | **Ref control** |
| --- | --- | --- | --- | --- | --- | --- | --- |
| 1 | CD3 | PerCP | SK7 | BD | 345766 | 1:25 | Cells |
| 2 | CD4 | V450 | L200 | BD | 560811 | 1:50 | Cells |
| 3 | CD8 | FITC | DK25 | Dako | F076501-2 | 1:25 | Cells |
| 4 | CD45RA | PE-Cy7 | L48 | BD | 337186 | 1:50 | Cells |
| 5 | CCR7 | BV711 | 150503 | BD | 566602 | 1:25 | Cells |
| 6 | CD69 | APC-H7 | FN50 | BD | 560737 | 1:50 | Cells |
| 7 | CD137 | PE | 4B4-1 | Miltenyi | 130-119-885 | 1:50 | Cells |
| 8 | OX40 | BV605 | L106 | BD | 745217 | 1:25 | Cells |
| 9 | LIVE/DEAD | BV510 | N/A | ThermoFisher | L34973 | 1:200 | Cells |

#### Supplementary Table 2. Panel of different monoclonal antibodies used for phenotypical analysis of SARS-CoV-2 S-specific B-cells.

|  | **Marker** | **Fluorochrome** | **Clone** | **Company** | **Catalogue** | **Dilution** | **Ref control** |
| --- | --- | --- | --- | --- | --- | --- | --- |
| 1 | CD20 | Pacific orange | HI47 | ThermoFisher | MHCD2030 | 20 | Cells |
| 2 | CD138 | BUV737 | MI15 | BD | 612834 | 50 | Beads |
| 3 | CD56 | BV510 | HCD56 | Biolegend | 318340 | 50 | Cells |
| 4 | CD10 | BV605 | HI 10 a | Biolegend | 312222 | 100 | Beads |
| 5 | CD27 | APC-R700 | M-T271 | BD | 565116 | 100 | Cells |
| 6 | CD38 | APC-Fire810 | HIT2 | Biolegend | 303550 | 100 | Cells |
| 7 | PD-1 | BV480 | EH12.1 | BD | 566112 | 100 | Beads |
| 8 | caspase-3 | V450 | C92-605 | BD | 560627 | 100 | Beads |
| 9 | CD3 | BV510 | OKT3 | Biolegend | 317332 | 100 | Cells |
| 10 | CD19 | BUV395 | HIB19 | BD | 740287 | 200 | Cells |
| 11 | CD21 | BUV805 | BLy4 | BD | 742008 | 200 | Cells |
| 12 | CD5 | BV750 | L17F12 | BD | 747090 | 200 | Cells |
| 13 | CD73 | AF647 | AD2 | Abcam | 243083 | 200 | Cells |
| 14 | CCR10 | BB515 | 1B5 | BD | 564769 | 200 | Cells |
| 15 | IRF4 | APC | REA201 | Miltenyi | 130-100-915 | 200 | Cells |
| 16 | HLA-DR | BUV496 | G46-6 | BD | 749866 | 200 | Cells |
| 17 | IgD | BUV563 | I-A6-2 | BD | 741394 | 400 | Cells |
| 18 | IgM | BV570 | MHM-88 | Biolegend | 314517 | 400 | Cells |
| 19 | IgG | PE-CF594 | G18-145 | BD | 562538 | 400 | Cells |
| 20 | CD95 | PE-Cy5 | DX2 | Biolegend | 305610 | 400 | Cells |
| 21 | CD43 | PerCP-Cy5.5 | 1G10 | BD | 563521 | 400 | Cells |
| 22 | CD45RB | PE | MEM-55 | Biolegend | 310204 | 400 | Cells |
| 23 | CD11c | BV650 | Bu15 | Biolegend | 337237 | 400 | Cells |
| 24 | Ki67 | BV711 | Ki67 | Biolegend | 350515 | 400 | Cells |
| 25 | LIVE/DEAD | Blue |  | ThermoFisher | L23105 | 500 | Cells |
| 26 | CXCR3 | PE-Cy7 | G025H7 | Biolegend | 353719 | 1200 | Cells |
| 27 | IgA | APC-Vio770 | IS11-8E10 | Miltenyi | 130-113-999 | 1600 | Cells |

Supplementary Table 3. Reactogenicity in PLWH. Reactogenicity occurring within seven days after administration of the additional mRNA-1273 vaccination in PLWH. Values are number (percentage). AE: adverse events, NSAID: non-steroidal anti-inflammatory drug.

|  | | **Overall**  **N=62** |
| --- | --- | --- |
| **Any AE** | | 41 (66.1) |
| **Local AE** | | |
| Pain at the injection site | | 41 (66.1) |
| Redness at the injection site | | 10 (16.1) |
| **Systemic AE** | | |
| Generalised myalgia | | 12 (19.4) |
| Fever | | 4 (6.5) |
| Headache | | 9 (14.5) |
| Rash other than injection site | | 3 (4.8) |
| **Medication use** | | |
| Any medication | | 7 (11.3) |
|  | Paracetamol | 5 (8.1) |
|  | NSAID | 2 (3.2) |
|  | Other | 0 |

### Supplementary Figures

Supplementary Figure 1. Gating strategy to detect SARS-CoV-2 S-specific T-cells. Lymphocytes were gated based on size (FSC-A) and granularity (SSC-A) and single cells were gated on basis of FSC-A and FSC-H. Next, LIVE CD3-expressing T-cells were selected and sub-divided into CD4 or CD8 expressing sub-sets. For both CD4 and CD8 T-cells memory sub-sets were gated based on CD45RA and CCR7 expression and naïve T-cells were excluded from further analysis. Within the memory T-cell population activation was determined based on the expression of OX40 with CD137 for CD4 cells and CD69 with CD137 for CD8 cells.

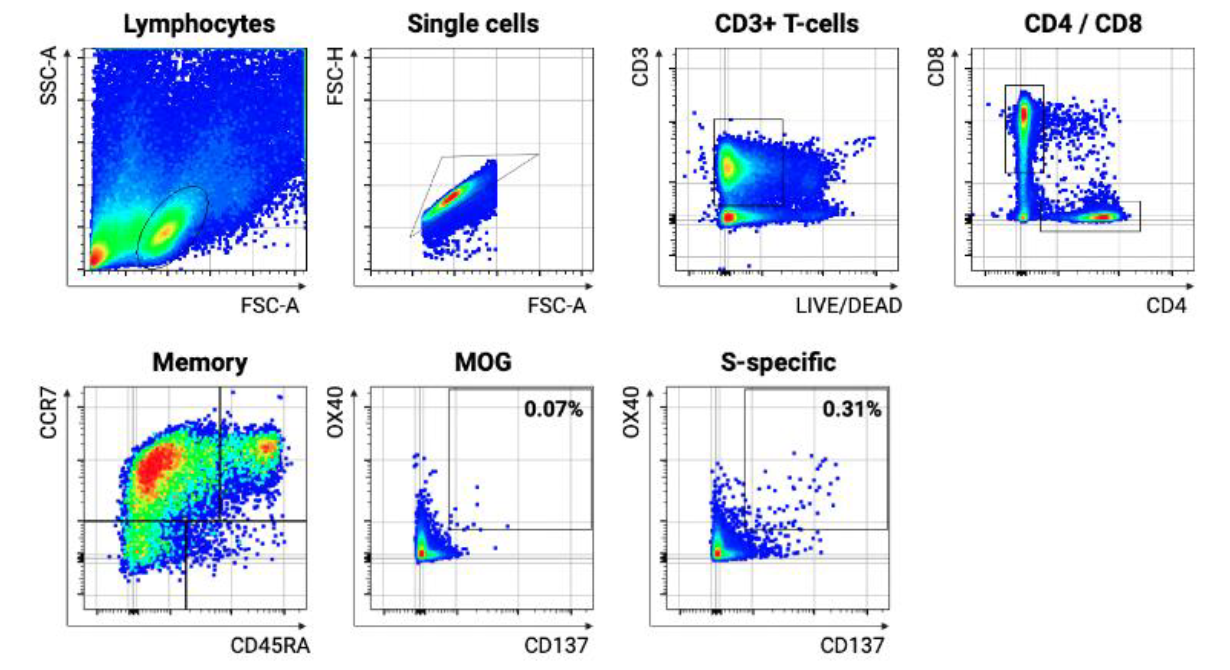

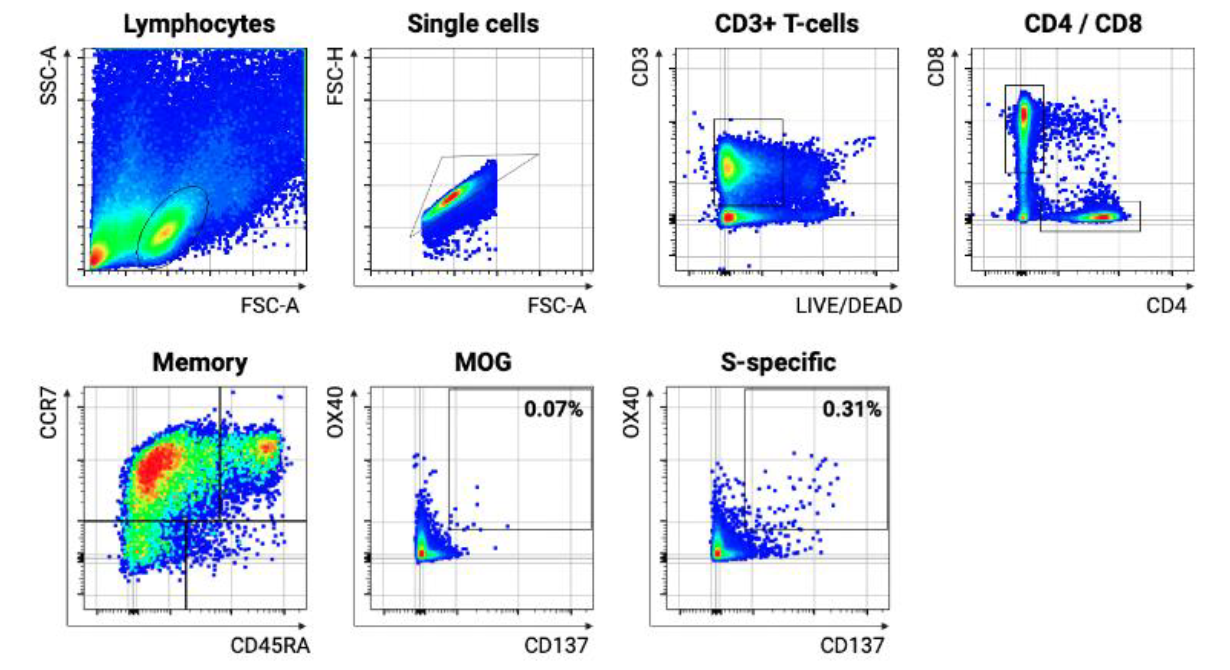

**DMSO**

Supplementary Figure 2. Gating strategy to detect SARS-CoV-2 S-specific B-cells. (A) Single-cell lymphocytes were first gated followed by exclusion of T and NK lymphocytes (CD3/CD56). Next, B-cells were gated (CD19) followed by exclusion of myeloid cells. B-cells were then exported. (B) B-cells were imported in OMIQ software and B-cells were excluded that were binding to streptavidin complexes that were not labeled with SARS-CoV-2 (decoy). SARS-CoV-2 S-specific B-cells were then gated based on double positivity for SARS-CoV-2 S-labeled tetramers (BUV661 and BUV615). Gated SARS-CoV-2 S-specific B-cells are shown for representative samples from T0 and T1 timepoints.

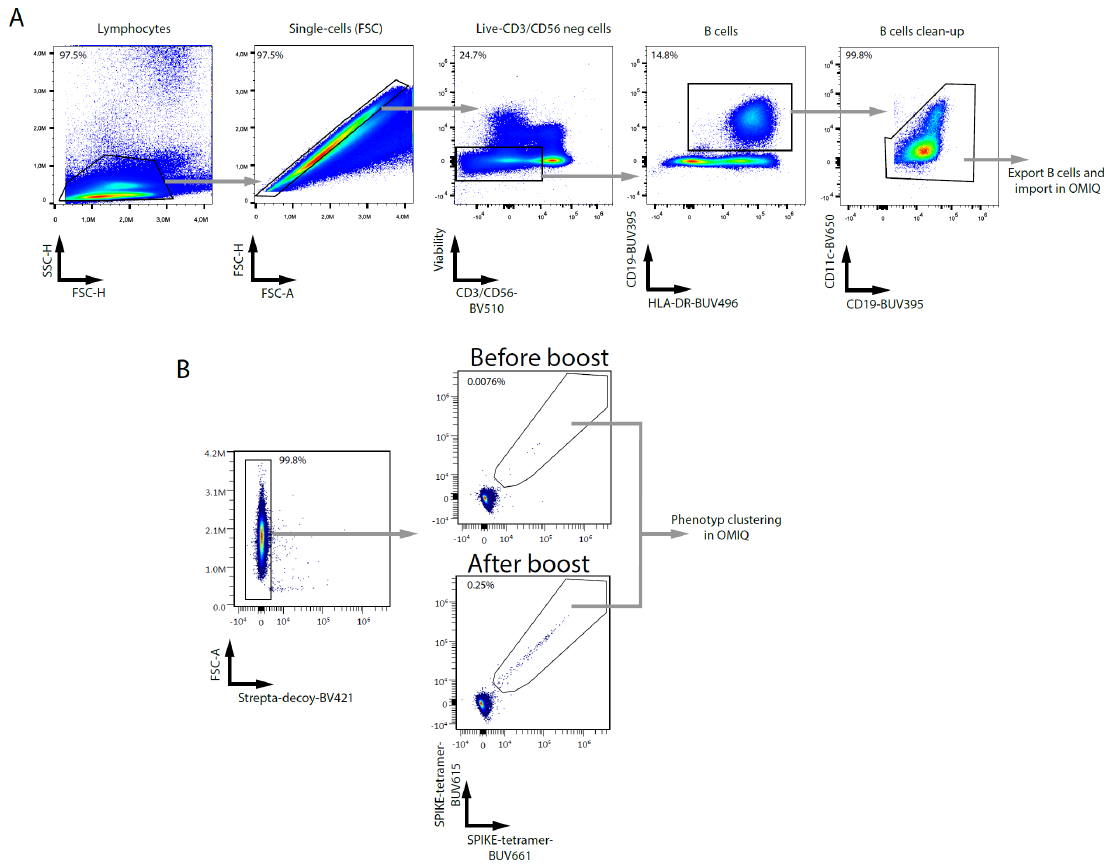

Supplementary Figure 3. Class-switching of SARS-CoV-2 S-specific B-cells. Isotype switching of B-cells was investigated by gating on IgD, IgM, IgMD, IgG and IgA expressing B-cells. Shown are plots of all samples (T0 and T1) that were concatenated. Normalized expressions of IgG, IgA, IgM and IgD are shown. In total, 6004 SARS-CoV-2 S-specific B-cells are shown.

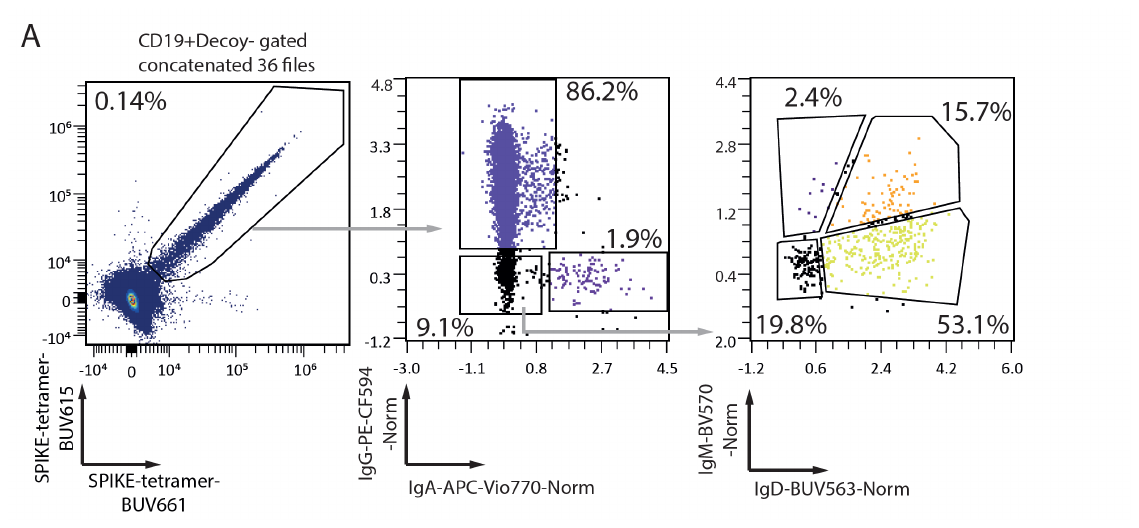

Supplementary Figure 4. Longitudinal analysis of SARS-CoV-2 S-specific B-cells after primary and additional vaccination in four PLWH. (A) Shown are the percentages of class-switched IgG+ SARS-CoV-2 S-specific B-cells at four different timepoints for four representative PLWH (A, B, C, D) with hyporesponse after the primary vaccination. (B) Isotype usage of SARS-CoV-2 S-specific B-cells are shown as stacked bars at AV1, AV2, T0, and T1 of the same four examples as panel A. Means and standard error of the means are shown in panel B. AV1: after primary vaccination timepoint 1, AV2: after primary vaccination timepoint 2, T0: before additional vaccination and T1: 28 days after additional vaccination.

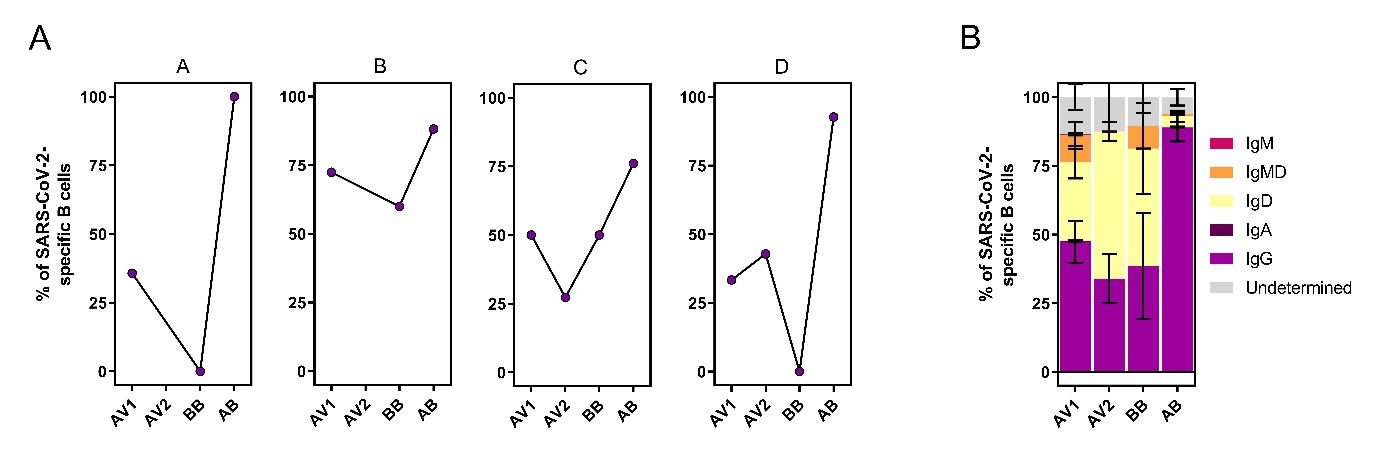

Supplementary Figure 5. Correlation of most recent CD4+ T-cell counts with percentage of IgG+ SARS-CoV-2 S-specific B-cells after additional vaccination. Percentages of IgG+ SARS-CoV-2 S-specific B-cells are shown as frequencies from total B-cells per individual. Each individual is colored according to the primary vaccination regimen (yellow: ChAdOx1-S and blue: BNT162b2). Pearson correlation analysis results on non-transformed data are depicted and linear regression results are shown as a black line with red shaded 95% Confidence Intervals.

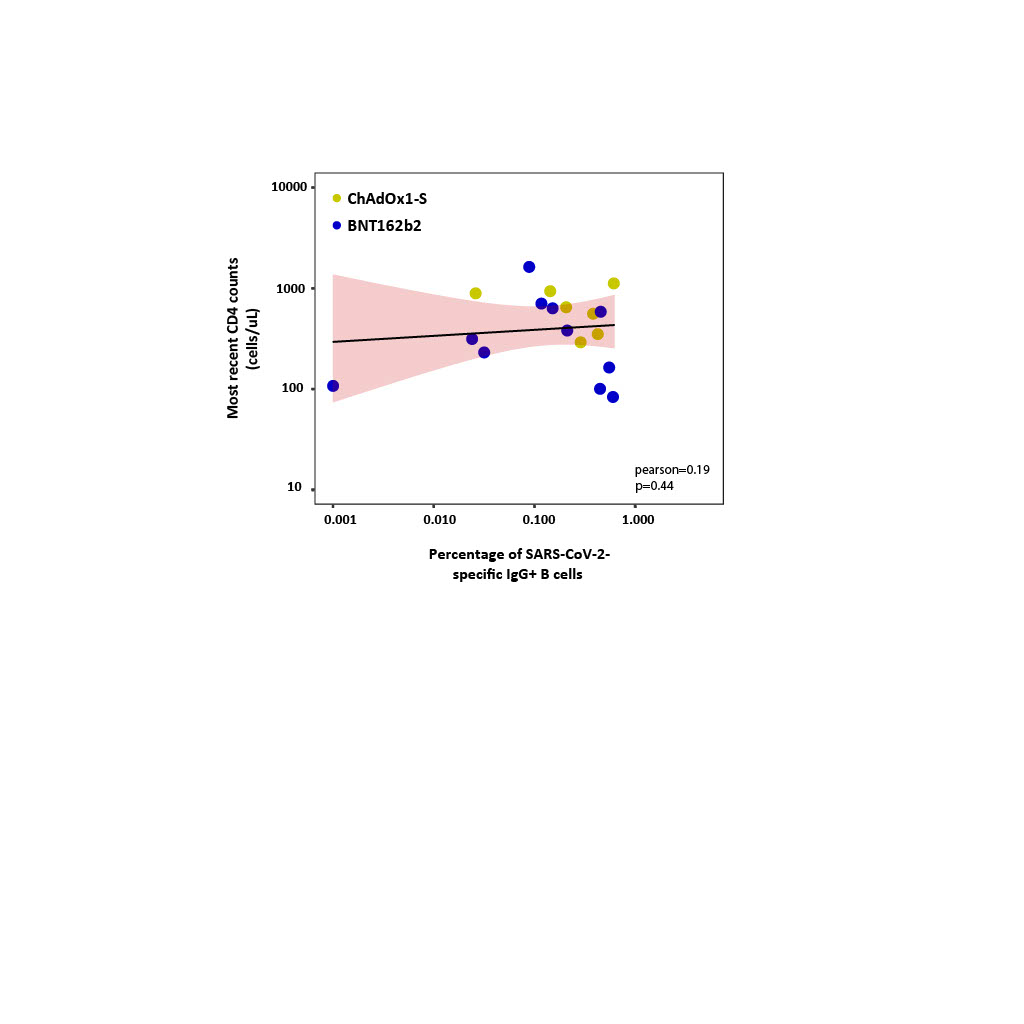

Supplementary Figure 6. Uniform Manifold Approximation and Projection (UMAP) of all SARS-CoV-2 S-specific B-cells. UMAPs of 6004 SARS-CoV-2 S-specific B-cells are shown and overlaid with the normalized expression of markers that were used to define the phenotype of SARS-CoV-2 S-specific B-cells, minus the markers shown in Figure 5D. The scale of the normalized expression is set min/max for each marker.

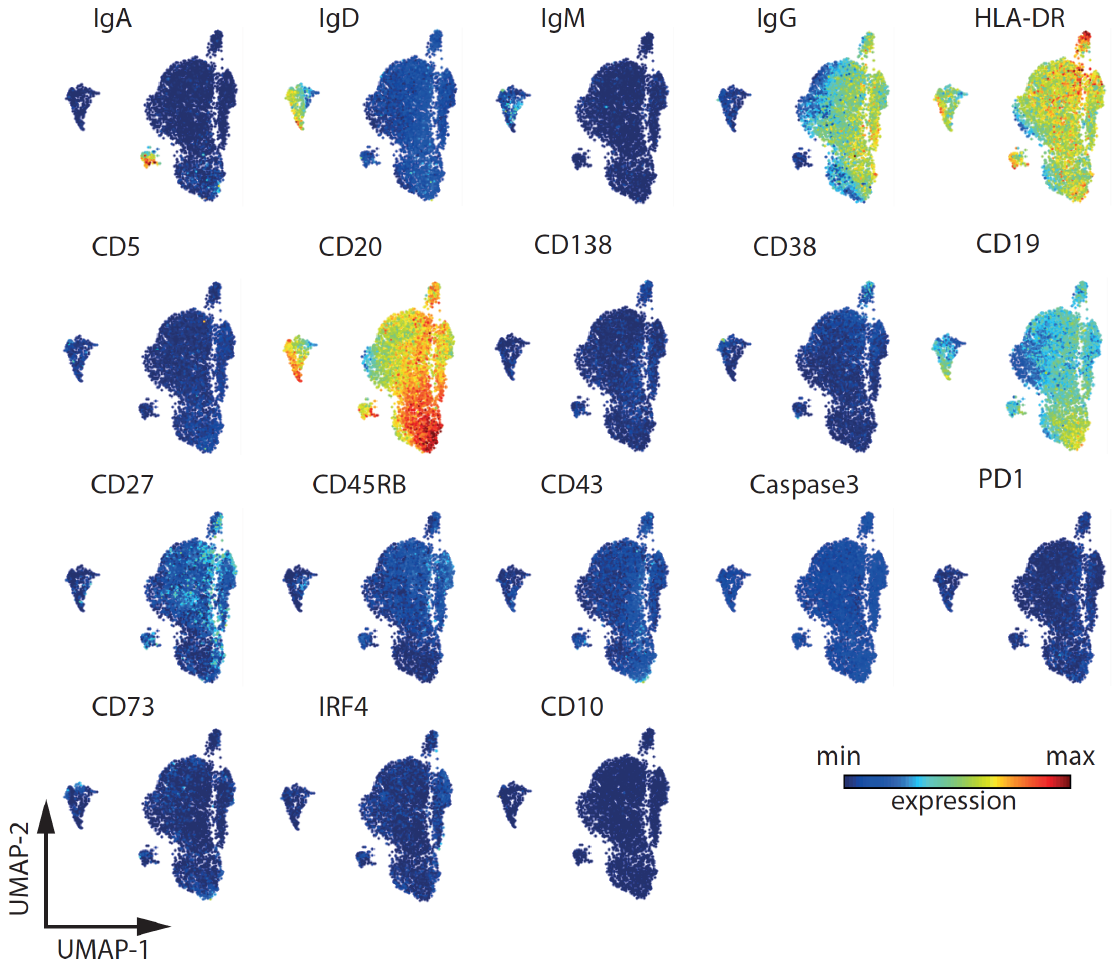
